# Complementary choroid plexus and locus coeruleus dysfunction in Parkinson’s disease progression

**DOI:** 10.64898/2026.07.02.26357179

**Authors:** Hongwei Li, Jia Jia, Jian Wang, Xiaoping Hu, Xiali Shao, Kai Liu, Jinhan Chen, Zhensen Chen, Lirong Jin, He Wang

**Affiliations:** Department of Radiology, University of Washington School of Medicine, Seattle, Washington, USA; Department of Rehabilitation Medicine, Longhua Hospital Affiliated to Shanghai University of Traditional Chinese Medicine, Shanghai, China; Department of Radiology, Zhongshan Hospital, Fudan University, Shanghai, China; Department of Bioengineering, University of California Riverside, Riverside, CA 92521, USA; Center for Advanced Neuroimaging, University of California Riverside, Riverside, CA 92521, USA; Institute of Science and Technology for Brain-inspired Intelligence, Fudan University, Shanghai, China; MOE Key Laboratory of Computational Neuroscience and Brain-Inspired Intelligence, Fudan University, Shanghai, China; Department of Neurology, Zhongshan Hospital, Fudan University, Shanghai, China; Department of Radiology, Shanghai Fourth People’s Hospital Affiliated to Tongji University School of Medicine, Shanghai, China

**Author notes:** These authors contributed equally to this work (Hongwei Li, Jia Jia, Jian Wang). **Correspondence to:** He Wang, Ph.D., Institute of Science and Technology for Brain-inspired Intelligence, Fudan University, Shanghai, China., Lirong Jin, Ph.D., Department of Neurology, Zhongshan Hospital, Fudan University, Shanghai, China, Zhensen Chen, Ph.D., Institute of Science and Technology for Brain-inspired Intelligence, Fudan University, Shanghai, China.

**Keywords:** Parkinson’s disease, choroid plexus, locus coeruleus, arterial spin labeling, neuromelanin-sensitive MRI, non-motor symptoms

## Abstract

Parkinson’s disease (PD) is a progressive neurodegenerative disorder commonly accompanied by cognitive decline, yet the mechanisms linking disrupted brain homeostasis to progressive cognitive impairment remain unclear. Emerging evidence suggests that the choroid plexus (ChP) and the locus coeruleus (LC) are involved in cerebrospinal fluid dynamics and norepinephrine regulation, respectively, but their longitudinal alterations and interrelationships in PD have not been systematically examined. We conducted a two-year longitudinal study including 90 PD patients and 51 healthy controls (HCs) undergoing multimodal MRI. ChP volume (ChP-V) was derived from T1-weighted structural imaging, ChP blood flow (ChP-BF) was assessed using pseudo-continuous arterial spin labeling, and LC integrity was ascertained with the contrast-to-noise ratio of the LC (LC-CNR) in neuromelanin-sensitive MRI. Group differences, longitudinal alterations, and associations with neuropsychological performance were examined. Over two years, PD patients showed progressive increases in ChP-V (F = 12.45, *p* < 0.001), reductions in ChP-BF (F = 18.98, *p* < 0.001), and declines in LC-CNR (F = 16.80, *p* < 0.001). Baseline LC-CNR was already reduced in PD compared with HCs (t = 3.023, *p* = 0.003). The longitudinal changes were more pronounced in male patients. ChP-BF was positively correlated with LC-CNR at baseline (*r* = 0.293, *p* = 0.006). Moreover, reductions in ChP-BF and LC-CNR were associated with worsening cognitive performance. While LC dysfunction was evident early in the disease course, progressive ChP alterations, particularly in ChP perfusion, provided additional information on longitudinal disease progression, supporting their combined value and highlighting the importance of gender-specific longitudinal monitoring.

## 1 | Introduction

Parkinson’s disease (PD) is a common progressive neurodegenerative disorder characterized primarily by motor symptoms but also by a wide range of non-motor manifestations that occur throughout the disease course^1^. Cognitive impairment is a frequent and debilitating non-motor feature that leads to a substantially reduced quality of life^2^. Although studies point to contributions from Lewy body and Alzheimer-type pathologies and from cholinergic, serotonergic, and noradrenergic dysfunction^3^, the mechanisms linking these pathological processes to disrupted brain homeostasis and PD-associated cognitive impairment are not yet fully understood.

Accumulating evidence has established the choroid plexus (ChP) as a critical regulator of cerebrospinal fluid (CSF) dynamics and neuroinflammation^4^, with both structural and functional alterations reported across aging^5^, neurodegeneration^6^, and psychiatric disorders^7^. These findings suggest that ChP dysfunction may also contribute to PD pathophysiology, potentially through impaired metabolic waste clearance or disrupted neuroimmune signaling. Consistent with this notion, magnetic resonance imaging (MRI) studies have demonstrated that increased ChP volume (ChP-V) is associated with reduced striatal dopaminergic uptake and elevated Unified PD Rating Scale (UPDRS-III) scores, suggesting a link between ChP alterations, dopaminergic degeneration, and motor impairment^8^. Moreover, ChP-V changes have also been reported in non-motor symptoms, including cognitive decline, possibly mediated by the pathological proteins in CSF, as reflected by the ratio between Aβ_1-42_ and α-synuclein (αsyn)^9^. However, most prior studies have focused on the morphological features of the ChP, whereas longitudinal changes in ChP volume and perfusion and their associations with cognitive decline in PD have rarely been examined.

Notably, a very recent cross-sectional study demonstrated that ChP perfusion exhibited distinct hemodynamic changes in PD and might play a mediating role in glymphatic system alterations^10^. This work indicated that simultaneous assessment of both structural and perfusion changes of the ChP in PD should be valuable, as these two dimensions capture distinct aspects of ChP and might interact with other imaging biomarkers to collectively contribute to the underlying glymphatic pathophysiology of PD.

A potential driving mechanism of CSF dynamics has been identified in a recent mouse study, which demonstrated that locus coeruleus (LC)-induced norepinephrine (NE) fluctuations drive the slow vasomotion, a primary force underlying CSF flow and thereby glymphatic clearance^11^, although this evidence was derived from sleep-state conditions. Another recent study in a rat model found that the CSF secretion rate was reduced when NE was delivered directly into the central ventricles and acted on the luminal side of the ChP^12^. Mammalian studies indicating that ChP function is influenced by NE concentrations can be traced back to the 1970s^13,14^, with the identification of a specific β-adrenergic-sensitive adenylate cyclase in the ChP providing biochemical evidence for this regulation^14^. Taken together, given that the LC is the major source of noradrenergic neurons in the brain and a region of particular relevance in PD, with established roles in cognitive and autonomic functions^15^, and in light of evidence linking LC degeneration to cognitive decline in PD^16^, we hypothesized that LC dysfunction may be indirectly involved in glymphatic function in PD and could offer insights distinct from those provided by ChP measures.

As advanced MRI sequences become more widely implemented and further optimized in clinical practice, the measurements described above have become increasingly accessible in clinical settings. Arterial spin labeling (ASL) is a noninvasive MR technique that quantifies cerebral blood flow (CBF) by magnetically inverting inflowing arterial spins. Pseudo-continuous ASL (PCASL) is now the recommended implementation^17^, which could provide highly reproducible and absolute quantification of ChP perfusion^18^. Neuromelanin, a by-product of NE metabolism, can be detected in vivo with neuromelanin-sensitive MRI (NM-MRI)^19^, which exploits the T1-shortening effects of paramagnetic neuromelanin to generate high-contrast images, with the LC appearing as a prominent high signal region in the brainstem^20^. In general, semi-quantitative metrics, such as contrast-to-noise ratio (CNR), are used to evaluate the LC^20^. These sequences are easy to implement and do not require the use of exogenous contrast agents, so they are particularly suitable for longitudinal clinical investigations.

Given the above considerations, in this longitudinal study, we aimed to investigate three MRI metrics: ChP-V, mean blood flow within the ChP (ChP-BF), and CNR of the LC (LC-CNR). Specifically, we first assessed group differences in these metrics between healthy controls (HCs) and PD patients at baseline (PD-V0) and at two-year follow-up (PD-V1). Second, we examined longitudinal changes in the three metrics within the PD group between V0 and V1. Third, given that ChP function might be modulated by NE, we explored the relationships between ChP-related metrics and LC-CNR. Finally, we investigated whether changes in these MRI metrics were associated with longitudinal changes in neuropsychological performance. Collectively, this study aims to elucidate the cross-sectional and longitudinal alterations of the ChP and the LC in PD, the relationships between these metrics, and their associations with disease progression, particularly cognitive decline.

## 2 | Materials and methods

### 2.1 | Participants

We recruited 90 PD patients and 51 HCs from the movement disorders clinic at Zhongshan Hospital. As previously detailed^21^, this study was approved by the ethical committee of Zhongshan Hospital, Fudan University. Written informed consent was obtained from all participants, prior to their enrollment in this longitudinal study. The diagnosis of PD was made according to the Movement Disorder Society (MDS) clinical diagnostic criteria for PD^22^ by two movement disorder specialists (J.L.R. and J.J.). Clinical evaluations were assessed and multi modal MRI was performed in PD patients at baseline (V0) and after 24 months (V1). Neuropsychology evaluations and MRI scans were also performed in control subjects at V0, the same as PD patients.

Based on our previous work^16^, Part III of the Unified Parkinson’s Disease Rating Scale (UPDRS-III) and Hoehn and Yahr (H&Y) staging were used for motor function assessment in patients with PD. Neuropsychological assessments were administered to evaluate multiple cognitive domains. Memory, visuospatial function, language, attention, and executive function were assessed using the following cognitive test: the Montreal Cognitive Assessment Basic (MoCA-B, representative general cognition), the Symbol Digit Modality Test (SDMT), Trail Making Test Parts A and B (TMT-A, TMT-B), the Stroop Color-Word Test (SCWT), the 30-item Boston Naming Test (BNT), the Animal Fluency Test (AFT), the Auditory Verbal Learning Test (AVLT), the Rey-Osterrieth Complex Figure Test (CFT), and the Clock Drawing Test (CDT). Depressive symptoms were evaluated with the 30-item Chinese version of the Geriatric Depression Scale (GDS)^23^. To screen for symptoms of rapid eye movement sleep behavior disorder (RBD), the Chinese version of the RBD Screening Questionnaire (RBDSQ) was employed. A score of ≥5 on the RBDSQ was used to indicate probable RBD according to a previous publication^24^. Furthermore, motor function was assessed using UPDRS-III and H&Y staging during the “OFF” medication state. In contrast, all neuropsychological evaluations and the GDS assessment were conducted during the “ON” medication state. The levodopa equivalent daily doses (LEDD) were calculated based on a commonly used standard^25^.

### 2.2 | MRI acquisition and postprocessing

All MR imaging was conducted on a 3-T MRI system (Discovery™ MR750, GE Healthcare, Milwaukee, WI, USA). A 3D high-resolution T1-weighted (T1W) structural imaging was acquired for ChP segmentation by using a brain volume imaging (BRAVO) sequence^26^ with the following parameters: repetition time/echo time/inversion time (TR/TE/TI) of 8.2/3.2/450 ms, field of view (FOV) of 240 mm, slice thickness of 1.0 mm with no gap, 136 slices, matrix size of 256×256, number of excitations (NEX) of 1, flip angle of 12°, and bandwidth of 31.25 kHz. The ASL images were acquired using a 3D PCASL sequence with background suppression and outward-direction spiral readout. The parameters of PCASL were as follows: TR/TE of 4830/10.5 ms, labeling duration/post labeling delay (PLD) of 1500/2500 ms, NEX of 3, flip angle of 155°, eight spiral arms with 512 points in each arm, and bandwidth of 62.5 kHz. NM-MRI data were acquired using a T1W fast spin-echo (FSE) sequence to calculate the LC-CNR with the following parameters: TR/TE of 600/13 ms, echo train length of 2, slice thickness of 2.5 mm with no gap, 16 slices, a matrix size of 512×320, FOV of 220 mm, NEX of 5. The axial slices were oriented parallel to the anterior commissure-posterior commissure (AC-PC) line, covering the region from the posterior commissure to the pons. Conventional MRI sequences were also obtained to rule out any other pathological findings that could potentially interfere with subsequent imaging analyses.

Automated segmentation of ChP in the lateral ventricles was first performed according to the Gaussian mixture model (GMM) segmentation method^27^. This approach has demonstrated superior accuracy compared with FreeSurfer^27^, which has been used in several previous studies for automatic ChP segmentation^6,28,29^, but it could still lead to some misclassified voxels and may fail to delineate the precise boundaries of the ChP (**Figure S1**). Therefore, one experienced neuroradiologist manually refined the segmentation masks using ITK-SNAP (version 3.8.0; http://www.itksnap.org/) to obtain the ChP volume (ChP-V) in milliliters (mL), and these masks were used for subsequent analyses. To assess inter-rater reliability, a second neuroradiologist independently performed the same refinement procedure, and a two-way random effects model was applied to calculate the intraclass correlation coefficient (ICC; see **Table S1** and **Figure S2** for details).

The voxel-wise cerebral blood flow (CBF) estimation was performed using the BASIL toolbox^30,31^, which is part of the FSL package^32^, with default settings following the recommendations of the ASL consensus paper^17^. The individual T1W images were also provided as inputs to BASIL, allowing the resulting CBF maps to be aligned and interpolated into the high resolution T1W space. The ChP masks obtained as described above were applied to the registered CBF maps to calculate the mean blood flow within the ChP masks (ChP-BF), expressed in units of mL/100□g/min.

The calculation of the CNR for the LC (LC-CNR) followed our previous work^16,33^, in which the LC was identified as the region with the highest signal intensity adjacent to the fourth ventricle on both sides. Circular regions of interest (ROIs) were placed in the bilateral LC and the pontine (PT), and the mean ± SD of the signal intensity within each ROI was recorded. The CNR of the LC was then defined as the mean signal difference between the LC and PT, divided by the SD of the PT signal^34^.

### 2.3 | Statistical analysis

All statistical analyses of demographic and clinical data were performed in MATLAB (R2020a, The MathWorks). Demographic characteristics, including age, gender, and education level, were first compared between the control and patient groups. Comparisons were performed using an independent two-sample t-test for age and educational level and Kolmogorov-Smirnov test for gender. The demographic characteristics of the PD group at V0 were further examined. Patients were first grouped by gender, and the Mann-Whitney U test was used to compare age, years of education, and RBDSQ between male and female patients. Next, patients were stratified according to the presence or absence of RBD. The Mann-Whitney U test was applied to compare age and education between the two groups, while the Kolmogorov-Smirnov test was used to compare the gender distribution. For the HCs, age and education were also compared between males and females using the Mann-Whitney U test.

Non-parametric tests were used for all clinical data. A Wilcoxon matched-pairs signed rank test was used to examine longitudinal changes between V0 and V1 within the PD group, and a Mann-Whitney U test was used to compare differences between the control and PD groups at both V0 and V1. Statistical significance was set at *p* < 0.05.

### 2.4 | Cross-sectional differences between PD and HCs

Normality and homogeneity of variance for ChP-V, ChP-BF and LC-CNR across different groups were checked using the Shapiro-Wilk test and Levene test. The Mann-Whitney U test or two-sample t-test was used to compare MRI metrics between PD and HCs, as appropriate. A two-way analysis of covariance (ANCOVA) was further conducted to examine the effects of group (PD vs. HCs) and gender (male vs. female) on MRI metrics at both V0 and V1, including the main effects of group and gender as well as their interaction term, while controlling for age, education and white matter hyperintensity (WMH) volumes.

Then, both the PD and HCs were divided into male and female subgroups. Due to the reduced sample size, only the Mann-Whitney U test was used to compare ChP-V, ChP-BF, and LC-CNR between the PD and HC groups. A one-way ANCOVA was subsequently applied to examine the effect of group (PD vs. HCs) within each gender subgroup at both V0 and V1, with age, education and WMH volumes included as covariates. An additional Mann-Whitney U test was performed to compare the MRI metrics between males and females within the HCs group.

For all Mann-Whitney U tests and two-sample t-tests, the resulting *p*-values were corrected for multiple comparisons using the false discovery rate (FDR) method, with FDR correction applied separately to analyses of ChP-related metrics and LC-CNR.

### 2.5 | Longitudinal analyses in the PD

To examine longitudinal changes in ChP-V, ChP-BF and LC-CNR within the PD group, a repeated measures analysis of variance (ANOVA) was performed, with time (V0 vs. V1) as the within-subject factor and gender and RBD status as between-subject factors. Age, follow-up interval, baseline UPDRS-III scores, baseline WMH volumes, and years of education were included as covariates. The analysis assessed the main effect of time to determine whether these MRI metrics changed over time in PD, and the interactions of time with gender and RBD status to examine whether longitudinal changes differed across PD subgroups. Tukey post-hoc tests were applied for pairwise comparisons when significant main or interaction effects were observed.

### 2.6 | Relationships between ChP-V, ChP-BF and LC-CNR

Partial Spearman correlation analyses were performed between ChP-related metrics and LC-CNR, controlling for age, gender, and WMH volume, separately in the HC, PD-V0, and PD-V1 groups. In addition, within the PD group, correlations between the longitudinal changes in LC-CNR and ChP-related metrics were also analyzed, controlling for follow-up interval, changes in WMH volume, and gender. All *p*-values from the partial correlation analyses were corrected for multiple comparisons using the FDR method.

A permutation test was followed for correlations that were significant in the PD group but not in HCs. To test whether the strength of association between two MRI metrics differed between HCs and PD, we compared the partial Spearman correlation coefficients computed separately within each group while adjusting for relevant covariates. To assess the statistical significance of the observed difference, a non-parametric permutation test with 5,000 iterations was used. In each iteration, group labels (HC vs. PD) were randomly permuted while preserving each subject’s paired MRI measures and covariates. Partial Spearman correlations were then recomputed within the permuted groups and the difference between the two coefficients recorded to form an empirical null distribution. The two-tailed *p*-value was computed as the proportion of permuted differences equal to or exceeding the absolute value of the observed difference. Statistical significance was set at *p* < 0.05.

To examine temporal precedence and potential bidirectional association among ChP-V, ChP-BF and LC-CNR in PD, cross-lagged panel analysis (*lavaan* package, v0.6.19) were performed using R 4.5.1. All variables were standardized (z-scored) prior to modeling. The first cross-lagged panel model (CLPM-1) included autoregressive paths for each variable from V0 to V1, and the cross-lagged effects which were defined as follows: ChP-BF at V0 predicting ChP-V and LC-CNR at V1; LC-CNR at V0 predicting ChP-V and ChP-BF at V1; and ChP-V at V0 predicting LC-CNR at V1. Note that the cross-lagged path from ChP-V at V0 to ChP-BF at V1 was assumed not to exist in the model. The second cross-lagged panel model (CLPM-2) excluded non-significant paths from CLPM-1 and included age, gender, and WMH volume as covariates.

### 2.7 | Associations between clinical data and MRI metrics

Bivariate Spearman correlation analyses were performed to examine the associations between longitudinal changes in MRI metrics (ChP-V, ChP-BF, and LC-CNR) and clinical measures (MMSE, SDMT, TMT-A, SCWT, TMT-B, BNT, AFT, AVLT, CFT, CDT, GDS, and UPDRS-III) within the PD group. Multiple linear regression analyses were subsequently conducted to further investigate the relationships that were significant in the preceding bivariate analyses, with gender, education, changes in age, UPDRS-III scores, GDS scores, and WMH volume included as covariates. A *p*-value < 0.05 was considered statistically significant.

## 3 | Results

### 3.1 | Demographic and Clinical Characteristics

The participants’ demographic and basic clinical characteristics are summarized in **Table 1**, with additional clinical information provided in **Table S2**. At V0, there were no significant differences in age, gender, or years of education between PD group and HCs. In addition, male and female PD patients did not differ significantly in age, years of education, or RBDSQ scores (**Table S3**). PD patients with and without RBD also showed no significant differences in age, gender, or education (**Table S4**). No significant differences in age or years of education were observed between male and female HCs (**Table S5**). All recruited patients with PD had mild disease severity with H&Y stages of 1 to 2. In addition, the PD patients showed significantly elevated UPDRS-III scores and poorer performance on cognitive tests.

**Table 1.** Clinical and MR features in PD patients at baseline (V0), at follow-up visit (V1), and healthy controls (HCs).

|  | HC<br>(n=51) | PD-V0<br>(n=90) | PD-V1<br>(n=90) | Statistic differences (*p* values) |  |
| --- | --- | --- | --- | --- | --- |
|  |  |  |  | PD-V0 vs. HCs | PD-V1 vs. PD-V0 |
| Age(y) | 64.12±7.47 | 63.99±7.82 | 66.22±7.84 | 0.924 | - |
| Gender(M/F) | 22/29 | 54/36 | 54/36 | 0.054 | - |
| Education(y) | 12.10±2.50 | 11.44±3.21 | 11.44±3.21 | 0.211 | - |
| H&Y |  | 1.49±0.56 | 1.82±0.70 | - | 0.001 |
| UPDRS III score |  | 18.7±8.25 | 22.63±10.1 | - | 0.005 |
| LEDD | - | 198.75±216.08 | 438.43±219.34 | - | <0.001 |
| **MR features** |  |  |  |  |  |
| ChP-V | 1.81±0.45 | 1.88±0.51 | 1.94±0.51 | 0.424 | <0.001 |
| ChP-BF | 44.79±8.99 | 44.03±10.3 | 41.42±9.91 | 0.662 | 0.002 |
| LC-CNR | 2.41±0.53 | 2.10±0.60 | 2.10±0.68 | 0.003 | 0.969 |

PD-V0 versus HCs using Mann-Whitney U test and longitudinal changes in PD group using Wilcoxon matched-pairs signed rank test for clinical data, except independent two-sample t-test for age and Kolmogorov-Smirnov test for gender. The two-sample t-test and paired t-test were used for MR features. Abbreviations: PD, Parkinson's disease; HCs, healthy controls; M, male; F, female; H&Y, Hoehn and Yahr stage; UPDRS-III, Unified Parkinson's Disease Rating Scale; LEDD, levodopa equivalent daily dose; ChP-V, choroid plexus volume; ChP-BF, mean blood flow within the choroid plexus; LC-CNR, contrast-to-noise ratio of the locus coeruleus.

### 3.2 | Cross-sectional group differences in MRI measures

NM-MRI data were missing for three control subjects and two PD patients at V1. One PD patient exhibited apparent blurring artifacts in the PCASL data at V0, thus the PCASL data from both time points for this patient were excluded from further analyses. The Shapiro-Wilk test and Levene test showed that ChP-V, ChP-BF and LC-CNR in each group followed normal distributions and had homogeneous variances (*p* > 0.05, **Table S6** and **S7**). **Figure 1** showed the ChP-V and ChP-BF of a representative PD patient at V0 and V1.

**Figure 1.**
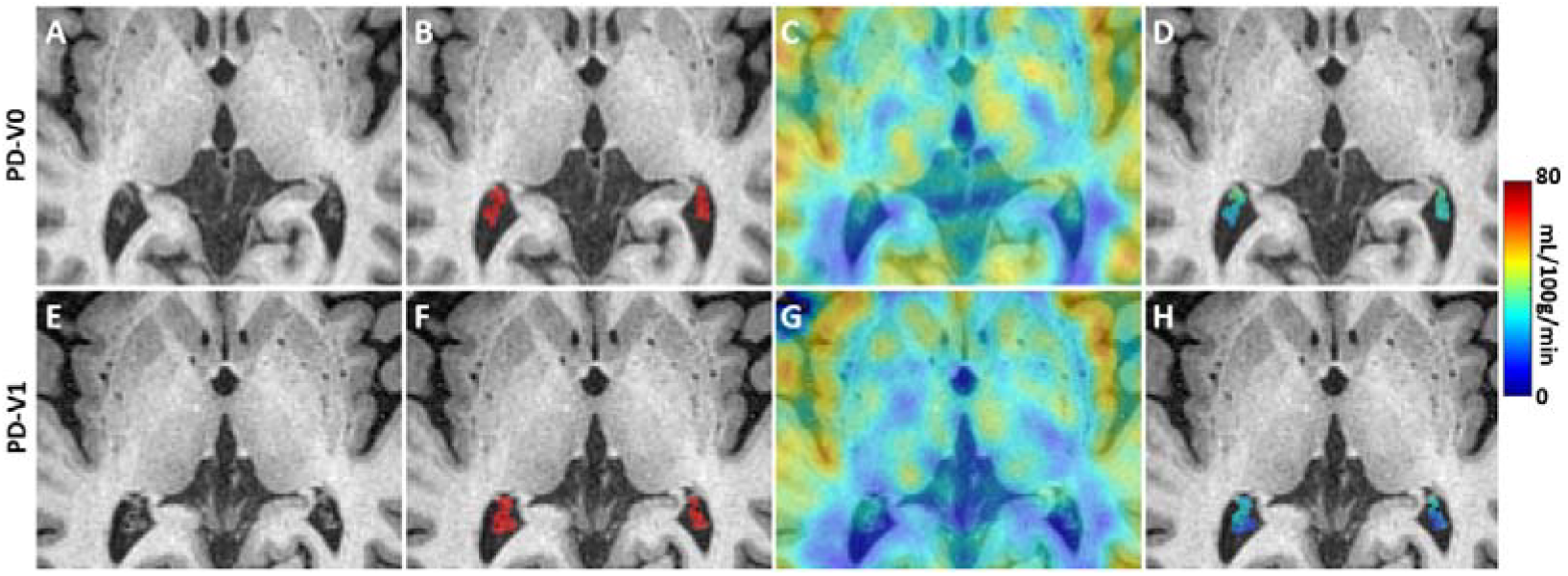
Choroid plexus (ChP) segmentation and cerebral blood flow (CBF) in a representative Parkinson’s disease (PD) patient at baseline (V0, top row) and follow-up (V1, bottom row). (A, E) T1W structural images. (B, F) ChP segmentation masks (red) overlaid on T1W images. ChP volume (ChP-V) was calculated as the sum of all voxel volumes within the mask. (C, G) CBF maps registered and overlaid onto T1W images. (D, H) Mean CBF within the ChP masks (ChP-BF), illustrating regional perfusion patterns within the ChP.

The two-sample t-tests showed no significant difference in ChP-V between PD and HCs at V0 (*p* = 0.424) or at V1 (*p* = 0.133). For ChP-BF, there was no significant difference between PD and HCs at V0 (*p* = 0.662) or at V1 (*p* = 0.048, *p*FDR = 0.095). The LC-CNR was significantly higher in HCs than in PD-V0 (t = 3.023, *p* = 0.003, *p*FDR = 0.009) and PD-V1 (t = 2.809, *p* = 0.006, *p*FDR = 0.009).

As for ChP-V, the two-way ANCOVA revealed a significant main effect of gender for both PD-V0 versus HCs (F = 18.40, *p* < 0.001) and PD-V1 versus HCs (F = 18.41, *p* < 0.001), as well as a significant group × gender interaction (F = 4.46, *p* = 0.036; F= 4.45, *p* = 0.037), respectively. The main effect of group was not significant for either comparison (PD-V0 vs. HCs: F = 0.04, *p* = 0.848; PD-V1 vs. HCs: F = 0.51, *p*= 0.476). With ChP-BF, only the significant main effect of gender was found for both PD-V0 versus HCs (F = 20.67, *p* < 0.001) and PD-V1 versus HCs (F = 21.29, *p* < 0.001). As for LC-CNR, the two-way ANCOVA revealed a significant main effect of group for both PD-V0 versus HCs (F = 6.90, *p* = 0.010) and PD-V1 versus HCs (F = 3.952, *p* = 0.048), and a significant main effect of gender (F = 12.90, *p* < 0.001; F = 27.47, *p* < 0.001), respectively, whereas no significant group × gender interaction was found (F = 0.20, *p* = 0.656 at V0; F = 2.77, *p* = 0.099 at V1).

After separating PD patients and HCs into male and female subgroups, the Mann-Whitney U test revealed that only the male PD patients at V1 showed a significant increase in ChP-V (*p* = 0.033, *p*FDR = 0.183) and a significant decrease in ChP-BF (*p* = 0.037, *p*FDR = 0.183) compared to HCs, although these results did not remain significant after FDR correction. In contrast, female PD patients did not show any significant ChP-related differences at either time point. Similarly, only the male PD patients at V1 showed a significant decrease in LC-CNR (*p* = 0.017, *p*FDR = 0.043) compared with HCs, whereas female PD patients did not show any LC-CNR differences at either time point. Further details are provided in **Table S8-10**. In addition, male and female HCs did not show significant ChP-V and ChP-BF differences (*p* = 0.337 and *p* = 0.082, respectively), but female HCs exhibited higher LC-CNR than males (*p* = 0.013, *p*FDR = 0.043).

The one-way ANCOVA revealed no ChP-related differences between PD patients and HCs in either male or female subgroups at any time point, whereas a significant decrease in LC-CNR was observed in male PD patients at V0 (F = 5.48, *p* = 0.022) and V1 (F = 7.26, *p* = 0.009), but not in female PD patients. Further details are provided in **Table S11**.

### 3.3 | MRI metrics longitudinal changes in PD

For ChP-V, repeated-measures ANOVA revealed a significant change over time (F = 12.45, *p* < 0.001). There was a significant main effect of gender (F = 20.24, *p* < 0.001) and a significant time × gender interaction (F = 19.88, *p* < 0.001). No significant main effect of RBD status (F = 0.32, *p* = 0.575) or time × RBD status interaction (F = 0.16, *p* = 0.687) was observed. The Tukey post-hoc tests further indicated that the ChP-V significantly increased from V0 to V1 in both male (*p* = 0.008) and female (*p* = 0.023) PD patients. Male PD patients exhibited significantly higher ChP-V than female PD patients at both V0 and V1 (*p* < 0.001 for both).

For ChP-BF, repeated-measures ANOVA revealed a significant change over time (F = 18.98, *p* < 0.001). There was a significant main effect of gender (F = 20.55, *p* < 0.001) and a significant time × gender interaction (F = 18.25, *p* < 0.001). The main effect of RBD status did not reach significance (F = 3.47, *p* = 0.066), but the time × RBD status interaction was significant (F = 4.16, *p* = 0.045). The Tukey post-hoc tests further indicated that the ChP-BF significantly decreased from V0 to V1 in both male (*p* = 0.015) and female (*p* = 0.043) PD patients. Male PD patients exhibited significantly lower ChP-BF than female PD patients at both V0 and V1 (*p* < 0.001 for both). Moreover, the ChP-BF also significantly decreased from V0 to V1 in PD patients with (*p* = 0.025) and without RBD (*p* = 0.027). No significant difference in ChP-BF was observed between PD patients with and without RBD at V0 (*p* = 0.171), but PD patients with RBD exhibited significantly lower ChP-BF at V1 (*p* = 0.045).

For LC-CNR, repeated-measures ANOVA revealed a significant change over time (F = 16.80, *p* < 0.001). There was a significant main effect of gender (F = 13.66, *p* < 0.001) and a significant time × gender interaction (F = 18.58, *p* < 0.001). The main effect of RBD status was significant (F = 6.18, *p* = 0.015), and the time × RBD status interaction was also significant (F = 4.61, *p* = 0.035). The Tukey post-hoc tests did not show significant longitudinal changes from V0 to V1 within either gender or RBD status subgroups. Male PD patients did not show a significant difference from female patients at V0 (*p* = 0.053), but exhibited significantly lower LC-CNR at V1 (*p* < 0.001). PD patients with RBD showed significantly lower values than those without RBD at both V0 (*p* = 0.039) and V1 (*p* = 0.035).

The distributions and longitudinal trends of ChP-related metrics and LC-CNR across subgroups with significant interaction effects identified in the above analyses are illustrated in **Figures 2 and 3**, respectively.

**Figure 2.**
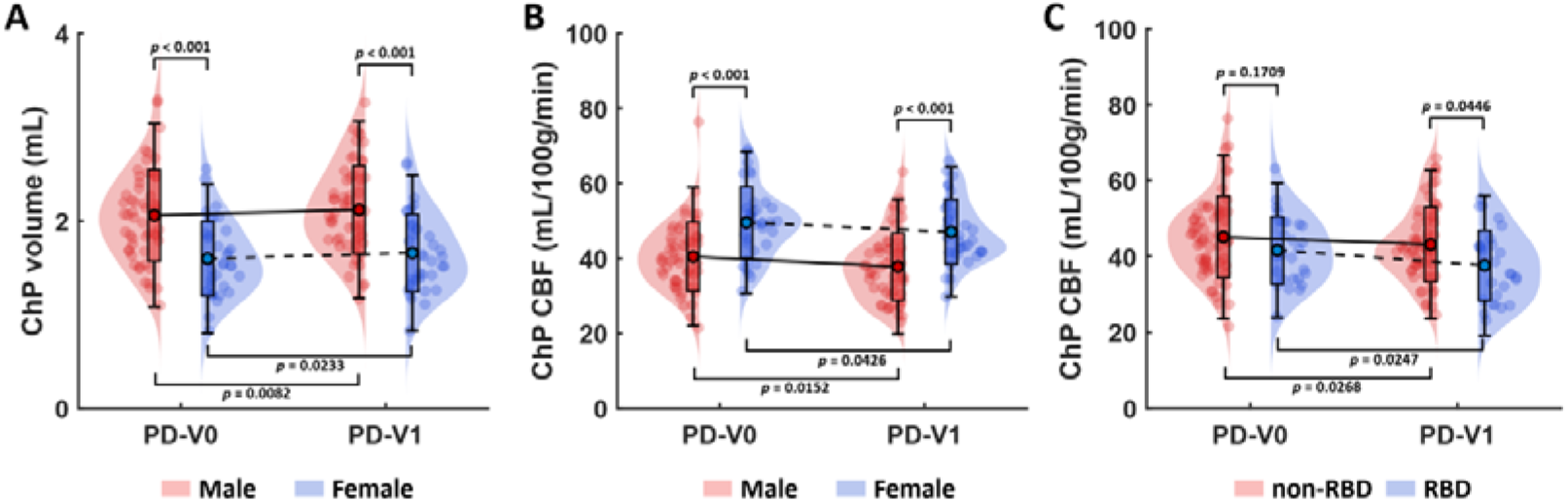
Longitudinal changes in choroid plexus (ChP) metrics across Parkinson’s disease (PD) subgroups. (A) Male ChP volume (ChP-V) increased from 2.06 ± 0.49 mL at V0 to 2.12 ± 0.47 mL at V1, while female ChP-V increased from 1.60 ± 0.40 mL at V0 to 1.65 ± 0.41 mL at V1. (B) Mean cerebral blood flow within the choroid plexus (ChP-BF) decreased from 40.44 ± 9.23 to 37.77 ± 8.99 mL/100g/min from V0 to V1 in male patients, and from 49.56 ± 9.47 to 47.06 ± 8.64 mL/100g/min in female patients. (C) PD patients without RBD showed a decrease in ChP-BF from 45.13 ± 10.78 to 43.17 ± 9.80 mL/100g/min from V0 to V1, while patients with RBD showed a decrease from 41.63 ± 8.88 to 37.62 ± 9.23 mL/100g/min. The *p*-values shown in the figure were obtained from repeated-measures ANOVA followed by Tukey post hoc tests. Error bars represent two standard deviations, and dots indicate the mean of the data distribution.

**Figure 3.**
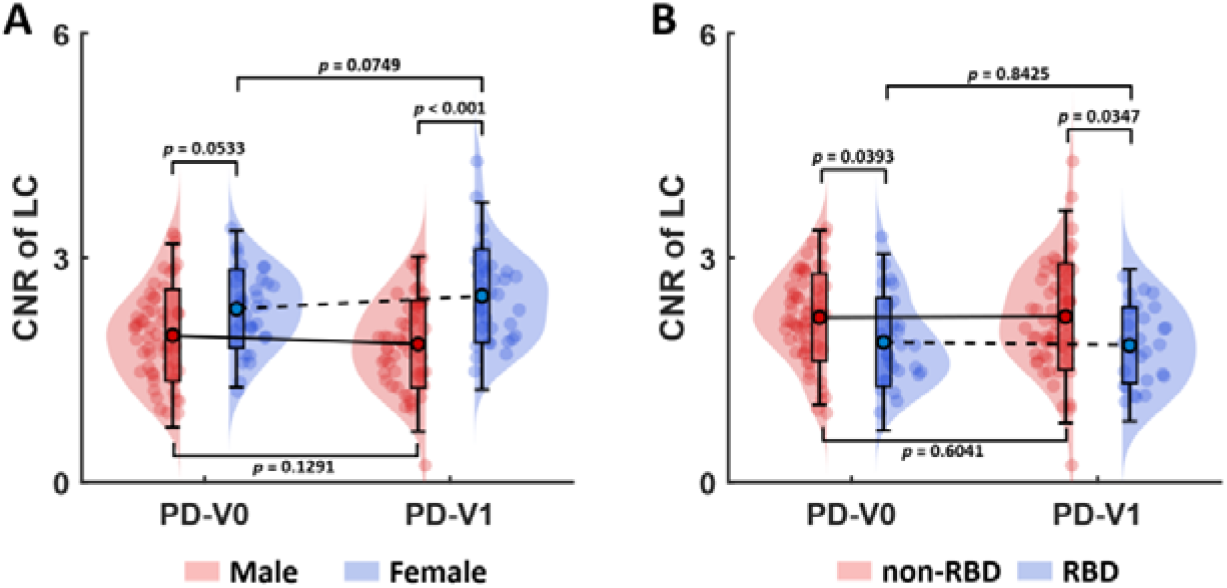
Longitudinal changes in the CNR for the LC (LC-CNR) across Parkinson’s disease (PD) subgroups. (A, B) No significant longitudinal changes from V0 to V1were observed within either gender or RBD status subgroups according to the Tukey post-hoc tests. At V1, male patients showed significant lower LC-CNR (1.85 ± 0.59) compared to female (2.49 ± 0.62). PD patients with RBD exhibited significantly lower LC-CNR at both V0 (1.87 ± 0.59) and V1 (1.83 ± 0.51) compared to patients without RBD at V0 (2.20 ± 0.58) and V1 (2.21 ± 0.71). The *p*-values shown in the figure were obtained from repeated-measures ANOVA followed by Tukey post hoc tests. Error bars represent two standard deviations, and dots indicate the mean of the data distribution.

### 3.4 | Associations among MRI metrics

A significant correlation between LC-CNR and ChP-related metrics was observed only in PD-V0, where LC-CNR was positively correlated with ChP-BF (*r* = 0.293, *p* = 0.006, *p*FDR = 0.037). No significant correlations were found in HCs or PD-V1. The permutation test showed that the positive correlation between LC-CNR and ChP-BF in PD-V0 was significantly different from that in HCs (*p* = 0.020). No significant correlations were found in the longitudinal changes of LC-CNR and ChP-related metrics in PD group. More details can be found in **Table S12** and **S13**.

In CLPM-1, the autoregressive effects from V0 to V1 were significant for all MRI metrics (all *p* < 0.001), and a significant cross-lagged effect was observed from ChP-V at V0 to LC-CNR at V1 (β = −0.183, *p* = 0.029). After including covariates in CLPM-2, the autoregressive effects remained significant for all metrics (all *p* < 0.001), whereas no cross-lagged effects survived. Model fit indices indicated excellent fit for both CLPM models. The detailed CLPM results are provided in **Table S14**, and the fit indices for the two CLPM models are shown in **Table S15**.

### 3.5 | Clinical data and MRI metrics correlation in PD

The changes in ChP-V and ChP-BF were significantly correlated with changes in TMT-A in bivariate Spearman correlation (*r* = 0.239, *p* = 0.023; *r* = −0.241, *p* = 0.023), respectively. After regressing out covariates, only changes in ChP-BF remained negatively correlated with changes in TMT-A (β = −2.26, *p* = 0.027), as shown in **Figure 4a**. The changes in LC-CNR were significantly correlated with changes in AVLT and TMT-A (*r* = 0.244, *p* = 0.023; *r* = −0.275, *p* = 0.010), respectively. After regressing out covariates, only changes in LC-CNR remained positively correlated with changes in AVLT (β = 2.49, *p* = 0.015), as shown in **Figure 4b**.

**Figure 4.**
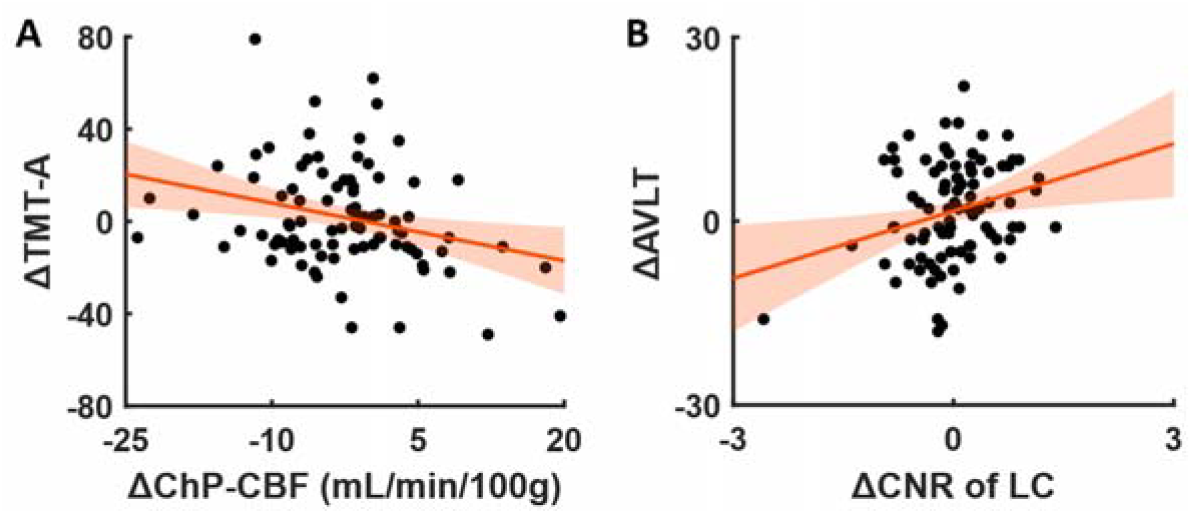
The correlation between longitudinal changes of MRI metrics and clinical data. (A) The prolonged competition time of Trail Making Test A (TMT-A) was negatively correlated with longitudinal blood flow reduction in choroid plexus (ChP).(B) A decline in Auditory Verbal Learning Test (AVLT) performance was associated with reduced CNR in the locus coeruleus (LC).

## 4 | Discussion

In this study, taking advantage of state-of-the-art non-invasive PCASL, high resolution T1W imaging and NM-MRI, the perfusion and volume of ChP, as well as LC integrity, were measured longitudinally in routine clinical settings. When considering the PD group as a whole, no significant differences in ChP-V or ChP-BF were found between PD patients and HCs, whereas LC-CNR was significantly reduced in PD patients at both baseline (V0) and two-year follow-up (V1). After stratification by gender, increased ChP-V, decreased ChP-BF, and significantly reduced LC-CNR were observed only in male PD patients at V1 (**Table S8-11**), relative to male HCs.

Within the PD group, longitudinal analyses demonstrated a significant increase in ChP-V and a significant decrease in ChP-BF from V0 to V1. Male patients exhibited significantly larger ChP-V and lower ChP-BF than female patients at both V0 and V1 (**Figure 2A, B**), whereas no such gender-specific differences were observed in HCs (**Section 3.2**). Although LC-CNR decreased significantly from V0 to V1 in the overall PD group, no significant longitudinal change was observed within gender-stratified subgroups. A significantly lower LC-CNR in male patients compared with females was observed only at V1 (**Figure 3A**), and a similar gender-specific trend was also present in HCs (**Section 3.2**). After separating the PD group according to RBD status, a significant group × time interaction was observed only for ChP-BF and LC-CNR. Both RBD and non-RBD patients showed longitudinal decreases in ChP-BF, but lower ChP-BF in RBD patients relative to non-RBD patients were only detected at V1 (**Figure 2C**). In contrast, LC-CNR did not show longitudinal changes in RBD or non-RBD patients, but significant differences between the two groups were evident at both V0 and V1. (**Figure 3B**). Furthermore, we also identified that longitudinal reductions in ChP-BF were correlated with greater increases in TMT-A completion time, and reductions in LC-CNR were associated with declines in AVLT performance, highlighting the clinical relevance of ChP and LC alterations to the progression of non-motor symptoms in PD.

Although the ChP has long been implicated in neurodegenerative disorders, most prior studies in PD focused primarily on morphological alterations, with inconsistent findings reported. Two studies based on the Parkinson’s Progression Markers Initiative (PPMI) database have reported divergent findings regarding ChP-V in PD. One study found no significant difference in ChP-V between PD patients and HCs^28^, whereas the other reported significantly lower ChP-V in PD compared with HCs^35^. In addition, two clinical studies conducted in Chinese PD cohorts with a disease duration of approximately 2-3 years both reported no significant difference in ChP-V between PD patients and HCs^36,37^, which is consistent with our findings. Significant ChP-V enlargement was observed in PD patients with freezing of gait compared with HCs, but longitudinal data from the PPMI cohort indicated that at baseline (approximately five years earlier), these patients did not differ significantly in ChP-V from PD patients without freezing of gait^36^. Our two-year longitudinal analysis demonstrated a significant change toward ChP-V enlargement with disease progression after adjusting for age, a pattern that appears directionally consistent with a recent study in another independent Chinese PD cohort reporting larger ChP-V in patients with a disease duration of approximately four years compared with HCs^10^. These findings also suggest that inconsistencies across studies might arise from differences in PD cohort characteristics, including disease severity, medication status, baseline cognitive abilities, and other factors. Moreover, findings from other longitudinal studies indicated that larger ChP-V might be linked to an increased risk of future freezing of gait^8^ and a higher probability of dementia conversion in early-stage PD patients^38^, providing further evidence for the involvement of ChP morphological changes in disease progression.

Attention to the association between ChP-BF and CSF production has been reported as early as the 1990s^39,40^, but studies specifically investigating ChP-BF in PD still remain extremely limited. In our study, similar to ChP-V, no significant differences in ChP-BF were observed between PD patients and HCs, but ChP-BF showed a significant decline over the two-year disease progression. The above-mentioned Chinese PD cohort study with a disease duration of approximately four years observed significantly lower ChP-BF compared with HCs^10^. This inconsistency might be attributed to differences in disease duration. Distinct from ChP-V, ChP-BF appeared to capture longitudinal differences between PD patients with and without RBD, with RBD patients showing a significantly lower ChP-BF compared to non-RBD patients after two years, also suggesting an accelerated decline of ChP-BF in the RBD patients (**Figure 2C**). Furthermore, although longitudinal changes in both ChP-V and ChP-BF were initially associated with longitudinal changes in TMT-A, after adjusting for covariates, only the association of ChP-BF with TMT-A remained significant (**Figure 4A**). The positive correlations between ChP-BF and both net CSF flow through the aqueduct of Sylvius^41^ and water flow across the blood-CSF barrier (BCSFB)^42^ were found in other ASL-based studies. Given this close association with CSF circulation, ChP-BF, as a functional biomarker, might provide a more robust independent measure than ChP-V for reflecting the progression of non-motor symptoms in PD.

In contrast to the ChP-related metrics, LC-CNR was already significantly reduced in PD patients compared with HCs at baseline, indicating early impairment of LC integrity. Unlike the ChP measures, LC-CNR showed longitudinal change over the two-year follow-up, but this longitudinal trend became non-significant after subgroup stratification, even for RBD patients. This might be partly attributable to reduced statistical power associated with smaller sample sizes, but might also be related to the relatively short two-year follow-up period and the notation that LC integrity degeneration happens early in PD pathology^43^.

Based on our findings, LC-CNR indeed provided cross-sectional and longitudinal perspectives on PD that were distinct from those offered by ChP-related measures. Moreover, the longitudinal decrease in LC-CNR was associated with worsening AVLT performance, indicating that LC alterations might be linked to changes in verbal episodic memory in PD. A similar association between LC-CNR and episodic memory has also been observed in aging populations^44^, and a positron emission tomography (PET) study also reported that higher LC catecholamine synthesis capacity appeared to attenuate tau-related memory deficits^45^. In addition, a recent PET study demonstrated a significant positive correlation between LC-CNR and NE transporter availability^46^, which indicated that LC-CNR derived from NM-MRI could indirectly reflect noradrenergic vulnerability. Collectively, these findings further support the notion that the LC-NE system is essential for episodic memory, with effects that appear to extend beyond PD, and the relatively slow longitudinal changes in LC-CNR over two years might also support the role of NE as a potential mediator of the compensatory aspect of cognitive reserve^47^, contributing to the brain’s ability to withstand pathological processes.

In our study, one of the most notable observations was the presence of gender-related differences in ChP-V, ChP-BF, and LC-CNR among PD patients. From a cross-sectional perspective, alterations including ChP-V enlargement, reduced ChP-BF, and decreased LC-CNR were observed exclusively in male PD patients, whereas no such differences were detected in female patients. Moreover, longitudinal analyses further demonstrated that male PD patients exhibited larger ChP-V, lower ChP-BF, and lower LC-CNR than female PD patients at both baseline and the two-year follow-up. Other neuroimaging studies have also consistently revealed distinct patterns of brain structural and functional alterations between male and female PD patients. One PPMI study comparing 149 male and 83 female *de novo* PD patients matched for disease duration and severity reported that male patients exhibited more extensive brain atrophy and greater disruption of white matter connectivity than female patients^48^. A similar finding was also found in another study, in which male *de novo* PD patients showed cortical thinning and reduced brain volumes in several brain regions, including the hippocampus and thalamus, together with worse motor performance and global cognition^49^. A data-driven long-term PD progression model also based on PPMI data revealed that female patients exhibited a 21.6% significantly slower disease progression rate compared to males^50^. Another recent study based on a cohort of 802 PD patients reported that men not only had a higher incidence of PD but also experienced a more rapid worsening of both motor and non-motor symptoms^51^. Taken together, our findings and the above-mentioned studies suggest that gender-related differences in PD span multiple levels, ranging from CSF circulation and the LC-NE system to brain structural and network properties, underscoring the need for gender-stratified monitoring and therapeutic strategies in PD management.

A significant association between ChP-BF and LC-CNR was identified in PD patients at baseline, and permutation testing against HCs indicated that this association was specific to the PD group. This correlation was not observed at follow-up, which might be partly explained by the preceding discussion. ChP-BF might be more sensitive to longitudinal changes during PD progression, whereas LC-CNR might predominantly reflect PD early pathophysiological alterations. These two measures captured distinct physiological processes, and the relatively short follow-up duration might constrain the detection of longitudinal associations. After adjusting for all covariates, the CLPM analysis did not reveal any significant cross-lagged effects, while significant autoregressive effects were observed for ChP-V, ChP-BF, and LC-CNR individually, suggesting that changes in these measures during disease progression might be independent and the longitudinal change of one metric did not reliably predict changes in the others. These results might suggest the presence of more complex mechanisms involving the LC and ChP in PD. The ChP-related metrics primarily reflected CSF circulation rather than the function of glymphatic system. In contrast, the along the perivascular space (ALPS) index, as a potential marker of glymphatic function, appeared to mediate the relationship between LC-CNR and cognitive function in PD^52^, as well as between ChP-related metrics and PD disease severity^10^. Future studies need to incorporate the ALPS index and extend the follow-up duration to better investigate the relationships among these measures.

We did not detect directional associations between LC-CNR and ChP-related measures, which might also reflect methodological limitations. Hauglund NL et al. performed measurements during sleep and reported that LC-NE fluctuations were temporally coupled with glymphatic clearance^11^. This coupling was observed in time-series dynamics rather than as a simple correlation of absolute signal levels. In contrast, MRI in typical clinical protocols is unable to capture temporal information, and our scans were acquired in awake participants. NM-MRI provides an indirect measure of neuromelanin rather than a direct measure of NE concentration. Although we speculate that the absolute LC-CNR values may show some relationship with LC-NE fluctuations, NM-MRI measurements of the LC remain constrained by relatively limited SNR at conventional 3T field strengths. PET radiotracers such as [^18^F]NS12137^53^ or [^11^C]methylreboxetine^46^ could be used to assess NE transporter distribution in clinical settings, but their high cost and limited accessibility reduce their practicality for large-scale or longitudinal studies.

Several limitations should be acknowledged in our study. First, the sample size was relatively small, which might reduce statistical power, particularly when performing subgroup analyses. Second, time-matched follow-up data were not obtained from HCs, which might limit our ability to assess longitudinal changes in ChP, LC, and cognitive function independent of aging. Also, RBD status was not evaluated using more accurate methods, such as polysomnography. Furthermore, multi-PLD PCASL was not used in this study because this sequence was not available on our scanner, and consequently, a relatively long PLD of 2500 ms was used in our protocol. Of course, multi-PLD PCASL could provide more comprehensive hemodynamic information, such as arterial transit time (ATT). However, it requires more complex post-processing, and different processing methods might affect the ICCs of ChP-BF measurements^18^. Although Liu Z et al. reported that some voxels in the ChP might have ATT values exceeding 2000 ms, leading to an approximately 10% underestimation with single-PLD compared to multi-PLD PCASL, they also proved that single-PLD could demonstrate comparable ICCs for ChP-BF measurement^18^. Since our PLD was longer than 2000 ms, this measurement bias is unlikely to have a substantial impact on the findings of our longitudinal analyses. Finally, manual adjustments of the ChP segmentation masks were time-consuming, limiting the feasibility of ChP analyses in larger cohorts. Previous studies have often relied solely on FreeSurfer, but our results have indicated that even with GMM-based segmentation, perfect segmentation cannot be achieved. Jeong SH et al. reported an agreement of 0.793 between neuroradiologist-drawn masks and GMM segmentation^8^, but careful manual correction of the ChP masks could improve the agreement to 0.941 (**Figure S2**). A deep learning model trained on multicenter database across the lifespan could achieve better segmentation performance compared with GMM methods^54,55^. Fine-tuning such a model on an independent dataset before applying it might represent a promising strategy.

In conclusion, PD progression was characterized by gradual enlargement of ChP-V and reduction in ChP-BF, whereas impairment of LC integrity was already evident at earlier stages of the disease. These longitudinal trends were more pronounced in male patients. ChP-BF was particularly sensitive to longitudinal change in patients with RBD and was associated with LC integrity at baseline, and its longitudinal reduction was linked to progressive cognitive decline in PD. Our findings also suggested that ChP and LC dysfunction might reflect relatively distinct pathophysiological processes during disease progression, indicating that glymphatic function and the LC-NE system might capture complementary aspects of PD progression and hold a potential value as longitudinal biomarkers.

## Data Availability

All data produced in the present study are available upon reasonable request to the authors.

## Data availability

The data that support the findings of this study are available from the corresponding author upon reasonable request.

## Author contributions

L.H. wrote the manuscript, designed and performed the imaging, and conducted the primary statistical analysis. J.J. collected and analyzed the clinical data. W.J. and L.K. established sequence acquisition parameters. W.J. and S.X. acquired the MRI data and corrected ChP masks. C.J. performed the initial organization and preprocessing of the MRI data. H.X., C.Z., J.L., and W.H. revised the manuscript. J.L. and W.H. supervised the study design. J.L. oversaw the inclusion and exclusion of clinical patient data. W.H. and C.Z. provided funding support for this project. All authors have read and approved the final manuscript.

