## Supplementary Materials for "Complementary choroid plexus and locus coeruleus dysfunction in Parkinson’s disease progression"

**Figure S1**. The choroid plexus (ChP) segmentation results from a representative subject. The segmentation obtained using the Gaussian Mixture Model (second column) showed missing voxels within the ChP and inclusion of incorrect voxels outside the ventricles (second row). After manual refinement by neuroradiologists, the quality of ChP segmentation was visually improved (last two columns).

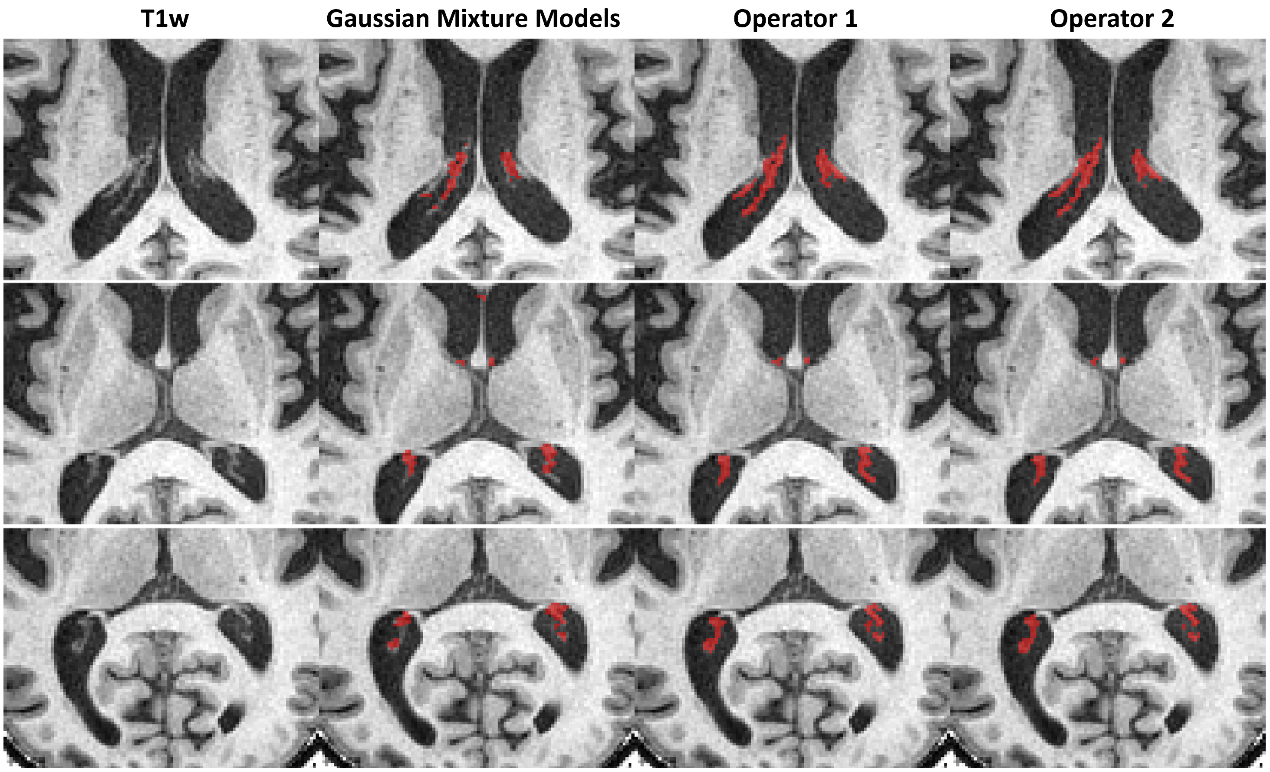

**Figure S2**. Spearman correlation between choroid plexus (ChP) volumes obtained by Gaussian Mixture Model (GMM) segmentation and manually delineated by two neuroradiologists in all included participants (51 healthy controls and 90 patients with measurements at two time points, 231 data samples). (A) The ChP volumes measured by operator 1 were significantly correlated with the GMM-based segmentation results (*r* = 0.511, *p* < 0.001), but several subjects exhibited pronounced outliers, which may be attributable to erroneous voxel inclusion in the automated GMM segmentation. (B) After manual refinement, the measurements between the two operators showed substantially improved consistency (*r* = 0.941, *p* < 0.001).

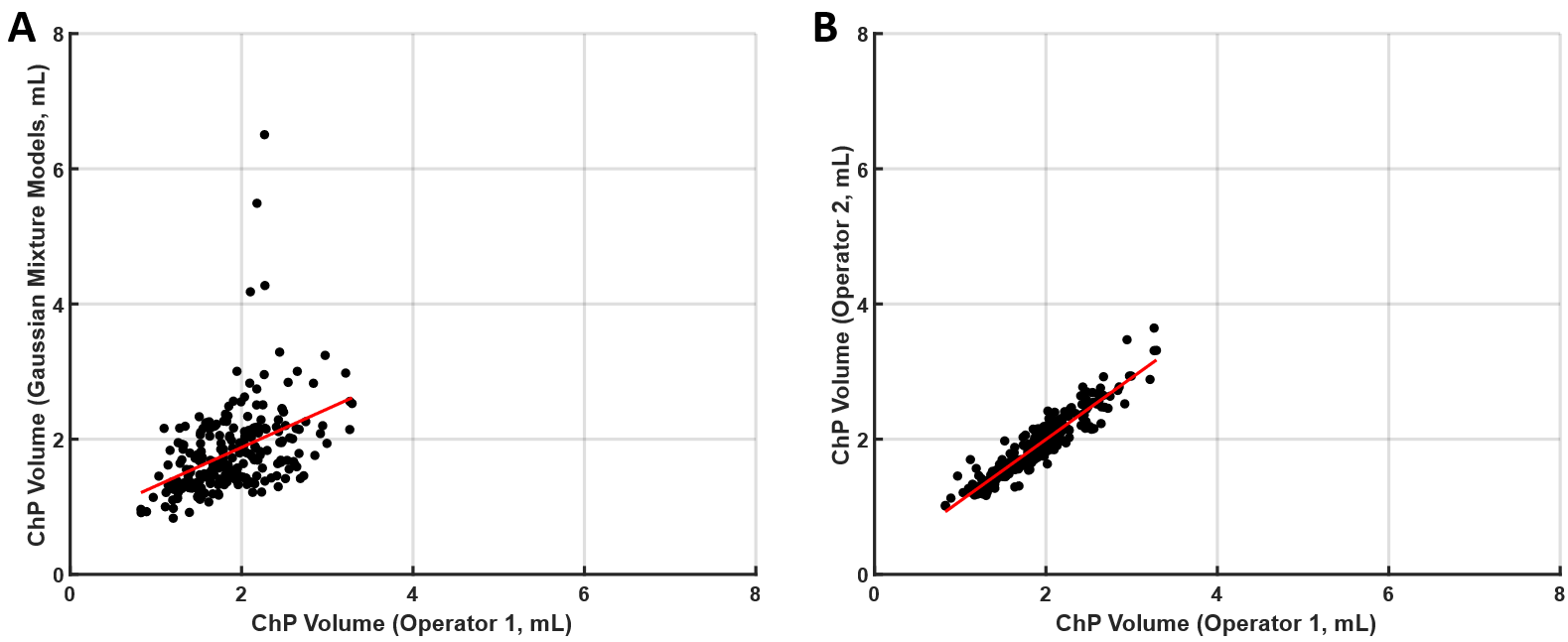

**Table S1**. Assessment of inter-rater reproducibility for choroid plexus volume (ChP-V) independently measured by two operators in all included participants (51 healthy controls and 90 patients with measurements at two time points, 231 data samples), along with the evaluation of measurement differences using the non-parametric Wilcoxon signed rank test.

| **MRI metrics** | **Intra-class coefficients of operators** | **Wilcoxon signed rank test *p* value** |
| --- | --- | --- |
| ChP-V | 0.93 (95%CI [0.91;0.95], *P* < 0.001 | 0.618 |

**Table S2**. Clinical features in PD patients at baseline (V0), at follow-up visit (V1), and healthy controls (HCs).

|  | HC  (n=51) | PD-V0  (n=90) | PD-V1  (n=90) | Statistic differences (*p* values) | |
| --- | --- | --- | --- | --- | --- |
|  |  |  |  | PD-V0 vs. HCs | PD-V1 vs. PD-V0 |
| GDS-30 scores | 6.43±5.22 | 9.00±6.30 | 11.54±5.61 | 0.015 | 0.005 |
| **Cognitive characteristics** | | | | | |
| MoCA-B | 26.80±1.83 | 25.51±2.31 | 25.19±3.22 | 0.001 | 0.461 |
| **Memory** | | | | | |
| AVLT-T | 31.24±8.06 | 27.39±9.64 | 28.94±11.19 | 0.017 | 0.319 |
| CFT-delay recall | 16.32±6.8 | 13.87±7.49 | 13.66±6.84 | 0.056 | 0.849 |
| **Visuospatial function** | | | | | |
| CFT copy | 32.78±2.71 | 31.29±4.29 | 31.48±3.63 | 0.027 | 0.750 |
| CDT | 9.33±0.79 | 8.97±1.34 | 10.07±9.06 | 0.077 | 0.256 |
| **Language** | | | | | |
| AFT | 18.82±5.71 | 17.90±5.23 | 17.09±5.39 | 0.331 | 0.307 |
| BNT | 25.76±2.93 | 24.78±3.29 | 24.63±3.8 | 0.077 | 0.785 |
| **Attention and working memory** | | | | | |
| SDMT | 44.86±10.89 | 36.02±10.97 | 33.68±11.77 | <0.001 | 0.001 |
| TMT-A | 51.86±18.59 | 69.00±26.34 | 71.10±32.26 | <0.001 | 0.633 |
| **Executive function** | | | | | |
| SCWT-C-time | 69.65±18.67 | 86.63±36.36 | 85.81±24.33 | 0.002 | 0.862 |
| SCWT-C-right | 47.74±2.32 | 46.59±3.85 | 46.04±3.64 | 0.057 | 0.331 |
| TMT-B | 119.22±41.48 | 152.45±43.16 | 166.57±56.68 | <0.001 | 0.016 |

PD-V0 versus HCs using Mann-Whitney U test and longitudinal changes in PD group using Wilcoxon matched-pairs signed rank test for clinical data. Abbreviations: PD, Parkinson’s disease; HCs, healthy controls; GDS, 30-item Chinese version of the Geriatric Depression Scale; MoCA-B, Montreal Cognitive Assessment Basic; AVLT, Auditory Verbal Learning Test; CFT, Rey-Osterrieth Complex Figure Test; CDT, Clock Drawing Test; AFT, Animal Fluency Test; BNT, 30-item Boston Naming Test; SDMT, Symbol Digit Modality Test; TMT-A, Trail Making Test A; TMT-B, Trail Making Test B; SCWT, Stroop Color-Word Test.

**Table S3**. Demographic characteristics and RBDSQ scores in male and female PD patients at V0. Comparisons were performed using the Mann-Whitney U test.

|  | **Male (n = 55)** | **Female (n = 35)** | ***p* values** |
| --- | --- | --- | --- |
| Age (y) | 65.31 ± 7.24 | 62.34 ± 8.25 | 0.067 |
| Education (y) | 11.78 ± 3.22 | 10.91 ± 3.16 | 0.153 |
| RBDSQ | 4.44 ± 3.35 | 3.43 ± 3.21 | 0.084 |

**Table S4**. Demographic characteristics in PD patients with and without RBD at V0. Age and education were compared using the Mann-Whitney U test, while the Kolmogorov-Smirnov test was applied to assess differences in gender distribution.

|  | **w/ RBD (n = 28)** | **w/o RBD (n = 62)** | ***p* values** |
| --- | --- | --- | --- |
| Age (y) | 66.60 ± 5.35 | 63.05 ± 8.41 | 0.110 |
| Education (y) | 11.68 ± 2.91 | 11.34 ± 3.35 | 0.548 |
| Gender (M/F) | 19/9 | 36/26 | 0.989 |

**Table S5**. Demographic characteristics in male and female HCs. Comparisons were performed using the Mann-Whitney U test.

|  | **Male (n = 22)** | **Female (n = 29)** | ***p* values** |
| --- | --- | --- | --- |
| Age (y) | 64.86 ± 7.62 | 63.55 ± 7.44 | 0.627 |
| Education (y) | 12.05 ± 3.00 | 12.14 ± 2.10 | 0.961 |

**Table S6**. Leven tests for the homogeneity of variance on ChP-V, ChP-BF, and LC-CNR in all included participants (51 healthy controls and 90 patients at V0 and V1) by performing ANOVA on the absolute deviations of the values from their group means.

|  | **ChP-V** | **ChP-BF** | **LC-CNR** |
| --- | --- | --- | --- |
| F values | 0.721 | 0.343 | 1.96 |
| df1 | 2 | 2 | 2 |
| df2 | 228 | 226 | 223 |
| *p* values | 0.487 | 0.710 | 0.144 |

**Table S7**. Normality tests of ChP-V, ChP-BF, and LC-CNR by Shapiro-Wilk test.

| **Group** | **ChP-V** | | **ChP-BF** | | **LC-CNR** | |
| --- | --- | --- | --- | --- | --- | --- |
|  | Statistic | *p* values | Statistic | *p* values | Statistic | *p* values |
| HCs | 0.974 | 0.284 | 0.988 | 0.889 | 0.983 | 0.699 |
| PD-V0 | 0.980 | 0.172 | 0.975 | 0.088 | 0.986 | 0.463 |
| PD-V1 | 0.985 | 0.402 | 0.976 | 0.104 | 0.982 | 0.224 |

**Table S8**. ChP-V differences between HCs and PD patients across male and female subgroups at both time points (mL). Due to the reduced sample size, comparisons were performed using the Mann-Whitney U test.

| **Group (M/F)** | **HCs (22/29)** | **PD-V0 (55/35)** | **PD-V1 (55/35)** | ***p* values** | ***p*FDR** |
| --- | --- | --- | --- | --- | --- |
| Male | 1.90 ± 0.46 | 2.06 ± 0.49 | 2.12 ± 0.47 | 0.124^a^, **0.033^b^** | 0.309^a^, 0.183^b^ |
| Female | 1.74 ± 0.43 | 1.60 ± 0.40 | 1.65 ± 0.41 | 0.205^a^, 0.403^b^ | 0.366^a^, 0.448^b^ |

^a^*p* values (HCs versus PD-V0); ^b^*p* values (HCs versus PD-V1).

**Table S9**. ChP-BF differences between HCs and PD patients across male and female subgroups at both time points (mL/100g/min). Due to the reduced sample size, comparisons were performed using the Mann-Whitney U test.

| **Group (M/F)** | **HCs (22/29)** | **PD-V0 (54/35)** | **PD-V1 (54/35)** | ***p* values** | ***p*FDR** |
| --- | --- | --- | --- | --- | --- |
| Male | 42.13 ± 7.91 | 40.44 ± 9.23 | 37.77 ± 8.99 | 0.369^a^, **0.037^b^** | 0.448^a^, 0.183^b^ |
| Female | 46.81 ± 9.38 | 49.56 ± 9.47 | 47.06 ± 8.64 | 0.220^a^, 0.946^b^ | 0.366^a^, 0.946^b^ |

^a^*p* values (HCs versus PD-V0); ^b^*p* values (HCs versus PD-V1).

**Table S10**. LC-CNR differences between HCs and PD patients across male and female subgroups at both time points. Due to the reduced sample size, comparisons were performed using the Mann-Whitney U test.

| **Group (M/F)** | **HCs (20/28)** | **PD-V0 (55/35)** | **PD-V1 (54/34)** | ***p* values** | ***p*FDR** |
| --- | --- | --- | --- | --- | --- |
| Male | 2.20 ± 0.44 | 1.96 ± 0.61 | 1.85 ± 0.59 | 0.055^a^, **0.017^b^** | 0.091^a^, **0.043^b^** |
| Female | 2.56 ± 0.54 | 2.32 ± 0.52 | 2.49 ± 0.62 | 0.090^a^, 0.432^b^ | 0.113^a^, 0.432^b^ |

^a^*p* values (HCs versus PD-V0); ^b^*p* values (HCs versus PD-V1).

**Table S11**. One-way ANCOVA results comparing PD and HCs within gender subgroups at V0 and V1, adjusting for age, education, and WMH volume.

|  | **Male** | | **Female** | |
| --- | --- | --- | --- | --- |
|  | F values | *p* values | F values | *p* values |
| ChP-V | 1.90^a^, 3.04^b^ | 0.172^a^, 0.085^b^ | 2.45^a^, 1.34^b^ | 0.123^a^, 0.251^b^ |
| ChP-BF | 0.857^a^, 2.91^b^ | 0.358^a^, 0.092^b^ | 1.69^a^, 0.022^b^ | 0.199^a^, 0.882^b^ |
| LC-CNR | 5.48^a^, 7.26^b^ | **0.022^a^**, **0.009^b^** | 2.76^a^, 1.34^b^ | 0.102^a^, 0.731^b^ |

^a^*p* values (HCs versus PD-V0); ^b^*p* values (HCs versus PD-V1).

**Table S12**. Partial Spearman correlation results for associations between ChP-related metrics and LC-CNR in HCs, PD-V0, and PD-V1 groups, adjusted for age, gender, and WMH volume.

|  | **LC-CNR** | | |
| --- | --- | --- | --- |
|  | HCs | PD-V0 | PD-V1 |
| **ChP-V** | *r* = -0.122,  *p* = 0.429, *p*FDR = 0.490 | *r* = -0.012,  *p* = 0.915, *p*FDR = 0.915 | *r* = -0.125,  *p* = 0.253, *p*FDR = 0.427 |
| **ChP-BF** | *r* = 0.163,  *p* = 0.292, *p*FDR = 0.427 | *r* = 0.293,  ***p* = 0.006, *p*FDR = 0.048** | *r* = 0.172,  *p* = 0.117, *p*FDR = 0.427 |

**Table S13**. Correlations between longitudinal changes in LC-CNR and ChP-related metrics in PD, adjusted for follow-up interval, WMH volume change, and gender.

|  | **Δ LC-CNR** |
| --- | --- |
| **Δ ChP-V** | *r* = -0.151, *p* = 0.169, *p*FDR = 0.427 |
| **Δ ChP-BF** | *r* = 0.110, *p* = 0.320, *p*FDR = 0.427 |

**Table S14.** Estimates from two cross-lagged panel model (CLPM) between LC-CNR and ChP-related metrics in PD.

|  | **CLPM-1** | | **CLPM-2** | |
| --- | --- | --- | --- | --- |
|  | *β* values | *p* values | *β* values | *p* values |
| Autoregressive effects |  |  |  |  |
| V0 (ChP-V) 🡪 V1 (ChP-V) | 0.958 | **< 0.001** | 0.962 | **< 0.001** |
| V0 (ChP-BF) 🡪 V1 (ChP-BF) | 0.709 | **< 0.001** | 0.711 | **< 0.001** |
| V0 (LC-CNR) 🡪 V1 (LC-CNR) | 0.491 | **< 0.001** | 0.456 | **< 0.001** |
| Cross-lagged effects |  |  |  |  |
| V0 (ChP-BF) 🡪 V1 (ChP-V) | -0.019 | 0.506 | / | / |
| V0 (ChP-BF) 🡪 V1 (LC-CNR) | 0.154 | 0.086 | / | / |
| V0 (LC-CNR) 🡪 V1 (ChP-BF) | 0.033 | 0.670 | / | / |
| V0 (LC-CNR) 🡪 V1 (ChP-V) | -0.017 | 0.559 | / | / |
| V0 (ChP-V) 🡪 V1 (LC-CNR) | -0.183 | **0.029** | -0.062 | 0.488 |

**Table S15.** Fit indices of two CLPM models.

|  | χ^2^ | df | CFI | TLI | SRMR | RMSEA |
| --- | --- | --- | --- | --- | --- | --- |
| CLPM-1 | 0.138 | 1 | 1.000 | 1.030 | 0.007 | 0.000 |
| CLPM-2 | 14.528 | 11 | 0.990 | 0.981 | 0.039 | 0.060 |

Abbreviations: χ^2^, chi-square; df, degrees of freedom; CFI, comparative fit index; TLI, Tucker-Lewis index; SRMR, standardized root mean square residual; RMSEA, root mean square error of approximation.
